# Accessibility of Canadian COVID-19 Testing Locations for People with Disabilities During the Third Wave of the COVID-19 Pandemic

**DOI:** 10.1101/2021.11.30.21267076

**Authors:** Sara Rotenberg, Jane Cooper, Matthew B. Downer

## Abstract

**Background:** Canadians with disabilities make up nearly a quarter of the population yet face barriers in accessing information about COVID-19 testing accessibility across the country.

**Objective:** No known studies evaluate the online information about the accessibility of COVID-19 testing locations. This study aimed to understand the accessibility of COVID-19 testing sites in Canada based on online information in March 2021.

**Methods:** Key accessibility features were identified to evaluate COVID-19 testing websites information on accessibility and data were extracted from the website to simulate the user experience of booking a COVID-19 test.

**Results:** All provinces and territories provided minimal accessibility information on their COVID-19 testing websites, except for Ontario. Out of 170 testing locations in Ontario, few had information about accessibility, with only 8.2% listing at least 3 of the 5 key accessibility features measured on their websites.

**Conclusions:** This paper demonstrates that more than a year into the pandemic, there existed a clear lack of accessibility information for Canadian testing locations for people with disabilities.

## Introduction

COVID-19 has had a significant and disproportionate impact on people with disabilities. For instance, the risk of death from COVID-19 in England was 3.1 times greater for men with disabilities and 3.5 times greater for women with disabilities than for the non-disabled population through 2020 (Bosworth et al., 2021). The risk of contracting SARS-CoV-2 and adverse COVID-19 outcomes is also higher for people with disabilities due to factors including place of residence, in-person care requirements, and high rates of chronic health conditions. Other studies have highlighted the broad social impacts, such as decreased access to health and social care services, unemployment, and violence throughout the pandemic (Shakespeare et al., 2021).

Testing has been a critical component of the “find-test-trace-isolate” paradigm to reduce the spread of SARS-CoV-2 and protect at-risk populations, like people with disabilities. A key tenet of the Canadian testing strategy has been the rapid and widespread creation of testing centers across the country (Health Canada, 2021). However, little attention has been paid to how to increase accessibility of testing locations and programs for individuals with disabilities, despite the fact that people with disabilities make up 22% of the Canadian population (Statistics Canada, 2018), and many require support to access testing because of various accessibility requirements (Kamalakannan et al., 2021). Having information about accessibility online is a critical component of ensuring there are incentives to get tested, as these barriers will hamper an individual’s intentions, regardless of their desire to get tested. When applied to Fishbein and Ajzen’s Theory of Reasoned Action (Fishbein & Ajzen, 1980), we see that without information about accessibility online (subjective norms), there are perceived or real barriers that impact people with disabilities’ decisions to get tested for COVID-19.

Improving the accessibility of Canada’s testing locations is important from a legal and ethical perspective for both COVID-19 and future public health emergencies (*Accessible Canada Act*, 2019; *Accessibility for Ontarians with Disabilities Act*, 2005; Doyle, 2021). This study aimed to examine the state of testing accessibility in Canada for people with disabilities by examining provincial/territorial COVID-19 testing websites and assessing local locations where location-level data were available to determine the accessibility of testing. Ultimately, these insights are important for improving the accessibility of testing locations in the future, improving ongoing COVID-19 testing and vaccination locations, and fostering the design of better, more accessible health emergency preparedness and response efforts for people with disabilities.

## Methods

### Data Collection and Extraction

A three-phase approach was adopted to ascertain accessibility at Canadian COVID-19 testing locations. First, given the lack of uniform accessibility guidelines in Canada, the authors devised a framework outlining what an accessible testing centre would look like by combining key aspects of the testing journey with accessibility features for each phase. This was developed using multiple sources, including lived experience from the authors, guidelines from the Accessibility for Ontarians with Disabilities Act (*Accessibility for Ontarians with Disabilities Act*, 2005), the Americans with Disabilities Act (Institute for Human Centered Design, 2017), recent guidance developed specifically for accessible COVID-19 testing and vaccine locations (Employment and Social Development Canada, 2020; McKee et al., 2021), and an ethical framework for public health (Kass 2001). These sources highlight the key accessibility features, as well as legal requirements for accessibility, which allowed us to develop a robust framework on which to measure testing location accessibility. From this, we selected key indicators to serve as a proxy for perceived accessibility of testing locations. While this may not be fully representative of accessibility at testing locations, this process mimics how people with disabilities would learn about testing locations and their accessibility, ultimately impacting their decision of whether or not to get tested.

Next, we conducted a pan-Canadian scan of provincial and territorial testing websites on March 20, 2021. For each website, we noted accessibility information such as booking and testing formats, information regarding accessible entrances, American Sign Language (ASL) interpretation, and other critical accessibility features at the province-level or by location.

Next, any province/territory that provided location-specific accessibility information was identified, and websites of individual local testing locations (community labs and assessment centres for people with symptoms) were examined for location-specific accessibility practices where available on March 20, 2021 and checked again on March 28, 2021. For testing, any pharmacies offering asymptomatic testing were excluded. At the location-level, the following information was extracted from websites of individual testing locations that provided accessibility information: appointment booking (appointment only or walk-in), drive-through testing availability, contact telephone number provided, wheelchairs available on site, booking format (telephone vs web-based), wheelchair-accessible entrance stated, care partners permitted, different testing modalities available, and whether ASL/interpreters were available. We also tabulated the number of locations that included information on multiple accessibility-related practices (Includes multiple booking formats, wheelchair-accessible entrance, care partner permitted, multiple testing modalities, ASL interpreters available). These factors were selected because they were most widely available, most likely to be advertised on a website, and were seen as feasible interventions to enhance accessibility. To ensure inter-rater reliability, all provincial websites and 15 location-specific websites were independently rated and compared between reviewers to ensure identical information was extracted. For the two conflicts, these locations were re-checked by both reviewers. No external research (i.e. site visits, interviews, etc.) was conducted, as we aimed to simulate what it would be like to determine the accessibility of locations as a potential user. All data extraction was performed by three authors (SR, MBD, JC).

## Results

### Province/Territory Level Accessibility Information

The accessibility information provided on all provincial and territorial COVID-19 testing websites is outlined in Table 1. Of the thirteen provincial/territorial websites, nine provided no accessibility information on their website. Both Newfoundland and Labrador and Yukon had websites that only included ASL-related accessibility considerations. No websites had specific easy-to-read information. Nova Scotia’s testing website provided high-level accessibility information, including a detailed process description, explicitly stating support animals are permitted, and information on how to request assistance if required. Only Ontario provided both provincial-level and location-specific accessibility information, and, therefore, was the only province or territory included in the second phase of data extraction and analysis. Overall, Ontario public health units varied in the degree to which locations that fell under their jurisdiction provided multiple accessibility-related services.

**Table 1:**
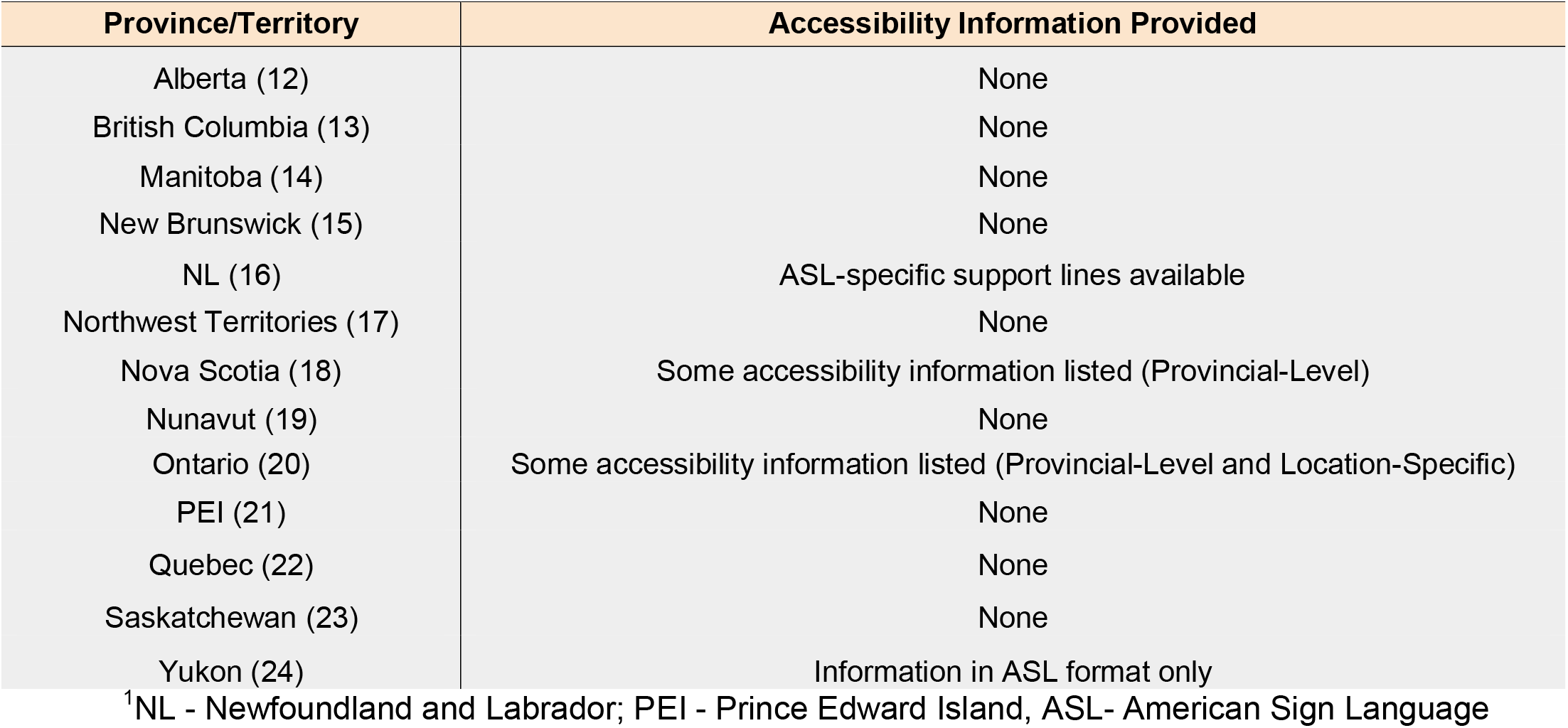
Accessibility Information Provided By Provincial Testing Websites (as of March 28, 2021)

## Discussion

This study demonstrates the limited information regarding accessibility of testing locations in Canada. At a national level, few provinces/territories provided any accessibility considerations in their respective testing websites, with most having no information on the accessibility for specific locations. Since only Ontario has location-level data for its assessment centres, it is difficult to both understand and measure the accessibility of testing locations from a national perspective. This has particular ramifications for people with disabilities across Canada, as inaccessible information and environments are significant barriers to equitable care (Kuper & Heydt, 2019). In particular, these data demonstrate how each step in the testing journey presents unique barriers to people with barrier to early access to treatment in this high-risk population. A year into the pandemic, it is deeply concerning that these gaps continue to exist for people with disabilities.

As **Table 2** suggests, limited accessible information or location-specific guidance at most people’s first point of entry—provincial websites—is a significant barrier to the decision and ability to seek testing. Phone numbers and multiple ways of booking are important and are disabilities. Although some people may find ways to access testing through their doctors or PHUs, each location’s website does not have information on accessibility, which can act as simple interventions. It is notable that almost all Ontario locations provided a phone number and half the locations allowed for multiple ways of booking. However, 47.1% of locations only allowed for online booking, which can be inaccessible to some people with disabilities. Considering most locations require an appointment time (85.9%), multiple booking modalities are important to ensure testing is universally accessible.

**Table 2:** Overview of Accessibility Information for Ontario Testing Locations (as of March 28, 2021)

| Accessibility Service | Number (%) of Locations Providing Service<br>(N=170) |
| --- | --- |
| **Appointment Booking** |  |
| *Appointment Only* | 146 (85.9%) |
| *No Info* | 24 (14.1%) |
| **Drive-Through Testing Used** |  |
| Yes | 41 (24.1%) |
| No | 129 (75.9%) |
| **Contact Telephone # Provided** |  |
| Yes | 157 (92.4%) |
| No | 12 (7.1%) |
| No Info | 1 (0.6%) |
| **Wheelchair Available On Site** |  |
| *No Info* | 170 (100%) |
| **Booking Format** |  |
| *Phone* | 67 (39.4%) |
| *Web* | 13 (7.6%) |
| *Phone or Web Options* | 85 (50.0%) |
| *Phone and Web Both Required* | 2 (1.2%) |
| *No Info* | 3 (1.8%) |
| **Wheelchair-accessible Entrance Stated** |  |
| Yes | 110 (64.7%) |
| *No Info* | 60 (35.3%) |
| **Care Partner Permitted** |  |
| Yes | 4 (2.4%) |
| No | 3 (1.8%) |
| *No Info* | 163 (95.9%) |
| **Different Testing Modalities Available** |  |
| Yes | 2 (1.2%) |
| No | 168 (98.8%) |
| **ASL/Interpreters Available** |  |
| Yes | 17 (10.0%) |
| *No Info* | 153 (90.0%) |
| **Multiple Accessibility Services<sup>1</sup>** |  |
| 0 | 33 (19.4%) |
| 1 | 70 (41.2%) |
| 2 | 55 (32.4%) |
| 3 | 14 (8.2%) |
| 4+ | 0 |

<sup>1</sup>Includes multiple booking formats, accessible entrance, care partner permitted, multiple testing modalities, interpreters available
<sup>2</sup>ASL: American Sign Language

Information about the location, testing method, and ability to bring care partners are also critical aspects of the decision and ability to seek testing. While drive-through testing may be more accessible to wheelchair users where accessible transport is provided, or people overwhelmed by sensory stimuli in new environments, it is important to note that this may be inaccessible to people with disabilities who are unable to drive (i.e. people with visual impairments or epilepsy). Thus, while it was good that several locations (24.1%) had this option, it should not be the only accessible solution. Conversely, for the testing modality, very few (1.2%) noted alternative testing modalities, such as saliva or swish-and-gargle formats, which may be more acceptable to people with sensory impairments (Thom & Turner, 2020), or ventilator users (SickKids Hospital, 2021). Finally, many people require care partners and/or service animals in order to have full access to environments. Little information on whether it was permitted to bring a care partner was available online; only 2.4% noted it was allowed to bring someone for support, and 1.8% actively discouraged it. While there are legitimate reasons for limiting the number of individuals at testing locations, it is important that these public health measures are balanced with access considerations for people who require this accommodation.

In terms of the accessible waiting line, approach, and entrance, we included several considerations. For instance, 64.7% of Ontario locations noted that they were physically accessible for wheelchair users or people who use mobility aids. This provides good insight into the accessibility of the location, but takes a narrow, binary definition of accessibility. For example, there was no information on wheelchairs availability; seating for those who cannot stand for long periods of time; or tactile guidance for people with visual impairments. Further, there was limited information about the inside of the facilities themselves, such as what the sensory environment would be like, which can deter people with sensory sensitivities, such as Autism, from seeking care (26). While our framework also notes the importance of having large signs; greeters; reduced sensory environments; clear masks and other methods of communication; strict public health measures, and other features, these were difficult to ascertain from provincial or location-specific websites.

In terms of the testing experience and providing results, limited information was publicly available online. While this does not necessarily indicate that locations are inaccessible, the lack of information may discourage individuals from seeking testing. For example, providing information on what to expect at each stage of the testing process and how results would be communicated, is helpful to ensure the individual understands what type of barriers may or may not exist in the testing environment. Outlining the procedure and public health measures are particularly important for people with disabilities who are at higher risk if they contract COVID-19 and/or who have difficulty waiting or being in overly stimulating sensory environments. Having more information available to walk people through the testing experience (videos, photos, text, etc.) can be helpful to ensuring access and removing barriers for those who need to get tested.

Moreover, communication is an integral component of the COVID-19 testing continuum. Not only are aids required to help people with sensory disabilities that impact communication, but assistance is also important for people who have difficulty comprehending information or responding to questions. As a result, locations should explain screening questions and testing procedures in multiple ways. Unfortunately, only 10.0% of Ontario testing locations stated that they have ASL interpreters available, and little to no information on other communication methods, such as availability of clear masks and clipboards

Overall, limited information was available on the accessibility of COVID-19 testing locations in Canada. While these results demonstrate several concrete gaps in testing location accessibility, these findings are also highly relevant to COVID-19 vaccination locations, which have become increasingly important as jurisdictions around the world continue to expand their COVID-19 vaccination programs (Rotenberg et al., 2021a). In addition, there is currently limited evidence or best-practices regarding how vaccination will be accessible. Findings from the present study can help inform how small changes can make existing locations accessible, as well as defining criteria for opening future, accessible locations.

Furthermore, the current inaccessibility of mainstream testing locations suggests it might be more feasible and accessible to formulate a twin-track approach to testing and vaccination locations. That is, not only trying to enhance the accessibility of communications, vaccination locations, and campaigns where possible to make them universally accessible, but also improving targeted interventions to reach all people with disabilities. This may include home visits for testing and vaccination, separate locations with trained staff, or specialized hours for those with disabilities, including slots with reduced sensory stimuli. This approach ensures that everyone with a disability has access to the critical testing and vaccination resources not only during COVID-19, but also beyond to ensure more equitable access to healthcare for people with disabilities.

### Knowledge Gaps and Future Directions

To the best of our knowledge, this is the first study evaluating the accessibility of COVID-19 testing locations for people with disabilities. Overall, there is limited evidence that people with disabilities and accessibility were adequately considered in the COVID-19 pandemic response across jurisdictions. In the future, studies should further examine how health environments--including those set up in pandemics and other health emergencies—are in-line with accessibility laws and health equity principles to ensure universal access for people with disabilities.

### Limitations

This study was exclusively conducted online, using only publicly available information. While this simulates the user’s experience of trying to find an accessible testing location and decide whether or not to get tested, it may not sufficiently capture the full accessibility or inaccessibility of a location. In addition, several locations noted that additional needs could be met in order to accommodate people with disabilities, but where these measures were not clear or publicly detailed online.

## Conclusion

Despite the fact that Canadians with disabilities make up 22% of the population (Government of Canada, 2018), and strong legal frameworks such as the Accessible Canada Act exist to ensure accessibility (*Accessible Canada Act, 2019*), the present work suggests COVID-19 testing locations across Canada fell short of providing information on universal accessibility. The framework we used to evaluate these recommendations highlights several key specific recommendations that would make each phase of the testing process accessible to people with disabilities, across impairment type (Rotenberg et al., 2021a). Many of these suggestions are low-cost or would require low effort to implement, as they are structural and behavioural changes, as opposed to physical ones. In particular, increasing the modalities in which information, booking, and results are available; clear guidance and support at the testing location; and more alternative options for people with disabilities (i.e., Home testing, alternative locations, etc.) would be helpful, immediate steps to improve access to testing. On the whole, these findings offer a key example of how a rapidly initiated COVID-19 testing program failed to center accessibility in their design and implementation. Moving forward, testing and vaccination programs around the world need to actively build accessible and universal design into all phases of their COVID-19 response, particularly testing and vaccination locations (Rotenberg et al., 2021a; Rotenberg et al., 2021b). The accessibility pillars and considerations set forth in this paper may be useful in designing and implementing universally accessible testing centers and preparing more robust, accessible plans for future emergencies and epidemics.

## Data Availability

All data produced are available online at the provincial public health websites.

## Acknowledgements

The authors acknowledge Drs. Yona Lunsky and Irfan Dhalla for their feedback and Tingting Yan for her assistance.

## Notes

**Funding and Conflicts:** SR and MBD receive funding from the Rhodes Trust. All authors declare no conflicts of interest.

### Competing Interest Statement

The authors have declared no competing interest.

### Funding Statement

SR and MBD receive funding from the Rhodes Trust. All authors declare no conflicts of interest.

